# Visualizing the invisible: The effect of asymptomatic transmission on the outbreak dynamics of COVID-19

**DOI:** 10.1101/2020.05.23.20111419

**Authors:** Mathias Peirlinck, Kevin Linka, Francisco Sahli Costabal, Jay Bhattacharya, Eran Bendavid, John P.A. Ioannidis, Ellen Kuhl

## Abstract

Understanding the outbreak dynamics of the COVID-19 pandemic has important implications for successful containment and mitigation strategies. Recent studies suggest that the population prevalence of SARS-CoV-2 antibodies, a proxy for the number of asymptomatic cases, could be an order of magnitude larger than expected from the number of reported symptomatic cases. Knowing the precise prevalence and contagiousness of asymptomatic transmission is critical to estimate the overall dimension and pandemic potential of COVID-19. However, at this stage, the effect of the asymptomatic population, its size, and its outbreak dynamics remain largely unknown. Here we use reported symptomatic case data in conjunction with antibody seroprevalence studies, a mathematical epidemiology model, and a Bayesian framework to infer the epidemiological characteristics of COVID-19. Our model computes, in real time, the time-varying contact rate of the outbreak, and projects the temporal evolution and credible intervals of the effective reproduction number and the symptomatic, asymptomatic, and recovered populations. Our study quantifies the sensitivity of the outbreak dynamics of COVID-19 to three parameters: the effective reproduction number, the ratio between the symptomatic and asymptomatic populations, and the infectious periods of both groups. For nine distinct locations, our model estimates the fraction of the population that has been infected and recovered by Jun 15, 2020 to 24.15% (95% CI: 20.48%-28.14%) for Heinsberg (NRW, Germany), 2.40% (95% CI: 2.09%-2.76%) for Ada County (ID, USA), 46.19% (95% CI: 45.81%-46.60%) for New York City (NY, USA), 11.26% (95% CI: 7.21%-16.03%) for Santa Clara County (CA, USA), 3.09% (95% CI: 2.27%-4.03%) for Denmark, 12.35% (95% CI: 10.03%-15.18%) for Geneva Canton (Switzerland), 5.24% (95% CI: 4.84%-5.70%) for the Netherlands, 1.53% (95% CI: 0.76%-2.62%) for Rio Grande do Sul (Brazil), and 5.32% (95% CI: 4.77%-5.93%) for Belgium. Our method traces the initial outbreak date in Santa Clara County back to January 20, 2020 (95% CI: December 29, 2019 - February 13, 2020). Our results could significantly change our understanding and management of the COVID-19 pandemic: A large asymptomatic population will make isolation, containment, and tracing of individual cases challenging. Instead, managing community transmission through increasing population awareness, promoting physical distancing, and encouraging behavioral changes could become more relevant.

## 1. Motivation

Since its outbreak in December 2019, the COVID-19 pandemic has rapidly swept across the globe and is now affecting 188 countries with more than 24 million cases reported worldwide [10]. In the early stages of a pandemic, doctors, researchers, and political decision makers mainly focus on symptomatic individuals that come for testing and address those who require the most urgent medical attention [15]. In the more advanced stages, the interest shifts towards mildly symptomatic and asymptomatic individuals who–by definition–are difficult to trace and likely to retain normal social and travel patterns [40]. In this manuscript, we collectively use the term “asymptomatic” for undetected individuals who either have mild symptoms that are not directly associated with COVID-19 or display no symptoms at all. Recent antibody seroprevalence studies suggests that the number of asymptomatic COVID-19 cases outnumbers the symptomatic cases by an order of magnitude or more [3, 4, 9, 12, 14, 18, 24, 32, 34, 46, 58, 59, 64, 65, 68, 66, 67, 71, 72, 74, 76, 78, 81]. Estimating the prevalence and contagiousness of these asymptomatic cases is critical since it will change our understanding of the overall dimension and the pandemic potential of COVID-19 [16]. Yet, at this stage, the effect of the asymptomatic population, its size, and its outbreak dynamics remain largely unknown [**?**].

The first evidence of asymptomatic individuals in a family cluster of three was reported in late January, where one individual was mildly symptomatic and two remained asymptomatic, with normal lymphocyte counts and chest computer tomography images, but positive quantitative reverse transcription polymerase chain reaction tests [53]. As of mid June, more than 50 studies have reported an asymptomatic population, twenty-three of them with a sample size of at least 500 [30], with a median undercount of 20 across all studies, suggesting that only one in twenty COVID-19 cases is noticed and reported. These studies are based on polymerase chain reaction or antibody seroprevalence tests in different subgroups of the population, at different locations, at different points in time [3, 7, 71]. To no surprise, the reported undercount varies hugely, ranging from 3.5 and 5.0 in Luxembourg [68] and Germany [71] to 543 and 627 in Iran [65] and Japan [9] respectively. Most of these studies are currently only available on preprint servers, but an increasing number is now passing peer review, including a study of 1402 individuals in Wuhan City with an undercount of 22.1 [81], a study of 400 health care workers in London with an undercount of 35.0 [77], a community spreading study of 131 patients with influenza-like symptoms in Los Angeles with an undercount of 100.0 [70], and a seroprevalence study in Los Angeles county with an undercount of 43.5 [69]. The reported trend across all studies is strikingly consistent: A much larger number of individuals displays antibody prevalence than we would expect from the reported symptomatic case numbers. Knowing the exact dimension of the asymptomatic population is critical for two reasons: first, to truly estimate the severity of the outbreak, e.g., hospitalization or mortality rates [16], and second, to reliably predict the success of surveillance and control strategies, e.g., contact tracing or vaccination [19].

While there is a pressing need to better understand the prevalence of asymptomatic transmission, it is also becoming increasingly clear that it will likely take a long time until we can, with full confidence, deliver reliable measurements of this asymptomatic group [6]. In the meantime, mathematical modeling can provide valuable insight into the tentative outbreak dynamics and outbreak control of COVID-19 for varying asymptomatic scenarios [40]. Many classical epidemiology models base their predictions on compartment models in which individuals pass through different stages as they experience the disease [33]. A popular model to simulate the outbreak dynamics of COVID-19 is the SEIR model [15], which is made up of four compartments for the susceptible, exposed, infectious, and recovered populations [2]. Here, to explicitly account for the asymptomatic population, we introduce an SEIIR model, which further divides the infectious population into symptomatic and asymptomatic groups [35]. Similar models have recently been used to study the general role of asymptomatic carriers in disease transmission [54] and to illustrate how asymptomatic individuals have facilitated the rapid spread of COVID-19 throughout China [40], South Korea [75], and Italy [20]. While it is tempting-and easily possible-to introduce many more sub-populations into the model, for example a pre-symptomatic, hospitalized, or mortality group [57], here, we focus on the simplest possible model that allows us to explore the role of the asymptomatic population throughout the COVID-19 pandemic. To systematically probe different scenarios, we combine this deterministic SEIIR model with a dynamic effective reproduction number, machine learning, and uncertainty quantification to learn the reproduction number-in real time-and quantify and propagate uncertainties in the symptomatic-to-asymptomatic ratio and in the initial exposed and infectious populations. We show that this not only allows us to visualize the dynamics and uncertainties of the dynamic contact rate, the effective reproduction number, and the symptomatic, asymptomatic, and recovered populations, but also to estimate the initial date of the outbreak.

## 2. Methods

### 2.1. Epidemiology modeling

#### SEIIR epidemiology model

We model the epidemiology of COVID-19 using an SEIIR model with five compartments, the susceptible, exposed, symptomatic infectious, asymptomatic infectious, and recovered populations. Figure 1 illustrates our SEIIR model, which is governed by a set of five ordinary differential equations,

**Figure 1:**
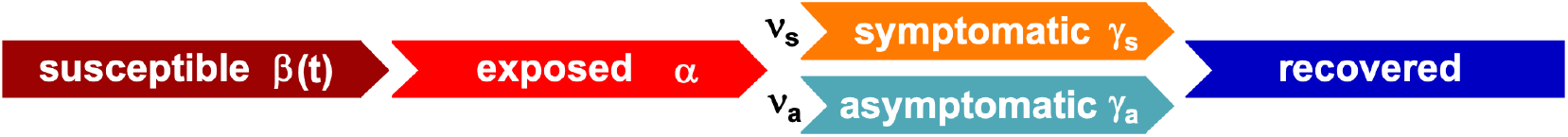
SEIIR epidemiology model. The SEIIR model contains five compartments for the susceptible, exposed, symptomatic infectious, asymptomatic infectious, and recovered populations. The transition rates between the compartments, *β*, *α*, and *γ* are inverses of the contact period *B =* 1/*β*, the latent period *A* = 1*/α*, and the infectious period *C =* 1/*γ*. The symptomatic and asymptomatic groups have the same latent period *A*, but they can have individual contact periods *B*_s_ = 1/β*_s_* and *B*_a_ = 1/*β*_a_ and individual infectious periods *C*_s_ = 1/*γ*_s_ and *C*_a_ = 1/*γ*_a_. The fractions of the symptomatic and asymptomatic subgroups of the infectious population are *v*_s_ and *v*_a_. We assume that the infection either goes through the symptomatic or the asymptomatic path, but not both for one individual.

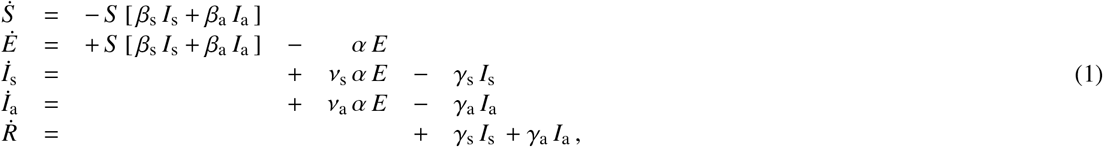

where the fractions of all five populations add up to one, *S + E + I_s_ + I_a_ + R =* 1. We assume that both the symptomatic group *I*_s_ and the asymptomatic group *I*_a_ can generate new infections. We introduce these two groups by fractions *v*_s_ and *v*_a_ of the subjects transferring from the exposed group *E*. We postulate that the two infectious groups *I*_s_ and *I*_a_ have the same latent period *A* = 1/*α*, but can have individual contact periods *B*_s_ = 1/*β*_s_ and *B*_a_ = 1/*β*_a_ to mimic their different community spreading, and individual infectious periods *C*_s_ = 1/*γ*_s_ and *C*_a_ = 1/*γ*_a_ to mimic their different likelihood of isolation. From the infectious fractions, we can derive the overall contact and infectious rates *β* and *γ* from their individual symptomatic and asymptomatic counterparts, *β*_s_, *β*_a_, *γ*_s_, and *γ*_a_,

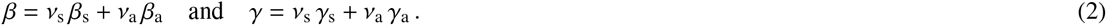

Similarly, we can express the overall contact and infectious periods *B* and *C* in terms of their symptomatic and asymptomatic counterparts, *B*_s_, *B*_a_, *C*_s_, and *C*_a_,

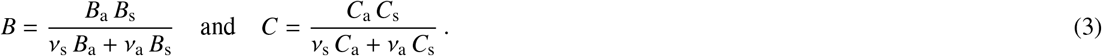

Naturally, the different dynamics for the symptomatic and asymptomatic groups also affect the basic reproduction number *R*_0_, the number of new infections caused by a single one individual in an otherwise uninfected, susceptible population,

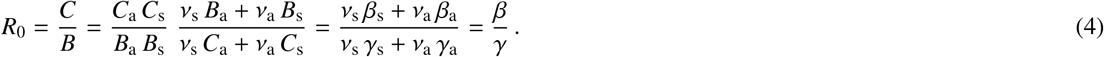

For a large asymptomatic group *v*_a_ → 1, the basic reproduction number approaches the ratio between the infectious and contact periods of the asymptomatic population, *R*_0_ → *C*_a_/*B*_a_, which could be significantly larger than the basic reproduction number for the symptomatic group, *R*_0_ = *C*_s_/*B*_s_, that we generally see reported in the literature. To characterize the effect of changes in social behavior and other interventions that may affect contact, we assume that the contact rate *β*(*t*) can vary as a function of time [43], but is the same for the symptomatic and asymptomatic groups,

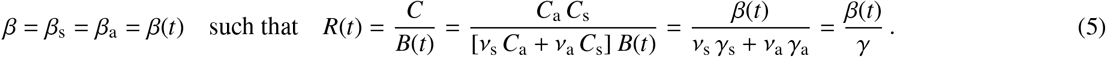

This introduces a time-varying effective reproduction number *R*(*t*), which is an important real time characteristic of the current outbreak dynamics.

#### SEIR epidemiology model

For the special case when the dynamics of the symptomatic and asymptomatic groups are similar, i.e., *β*_s_ = *β*_a_ = *β* and *γ*_s_ = *γ*_a_ = *γ*, we can translate the SEIIR model (1) into the classical SEIR model (6) with four compartments, the susceptible, exposed, infectious, and recovered populations [42, 44, 82, 83]. For this special case, we can back-calculate the symptomatic and asymptomatic groups from equation (6.3) as *I*_s_ = *v*_s_ *I* and *I*_a_ = *v*_a_ *I*. Figure 2 illustrates the SEIR model, which is governed by a set of four ordinary differential equations [28],

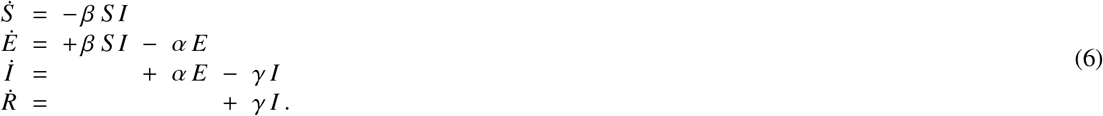

**Figure 2:**
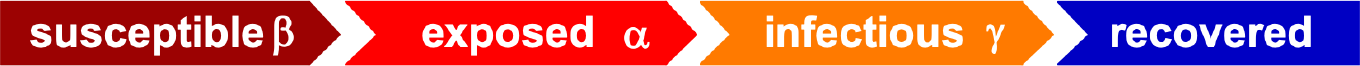
SEIR epidemiology model. The SEIR model contains four compartments for the susceptible, exposed, infectious, and recovered populations. The transition rates between the compartments, *β*, *α*, and *γ* are inverses of the contact period *B* = 1/*β*, the latent period *A* = 1/*α*, and the infectious period *C* = 1 /*γ*. If the transition rates are similar for the symptomatic and asymptomatic groups, the SEIIR model simplifies to the SEIR model with *I*_s_ = *v*_s_ *I* and *I*_a_ = *v*_a_*I*.

#### Discrete SEIIR epidemiology model

We focus on the SEIIR model with five ordinary differential equations (1), which we discretize in time using a finite difference approximation 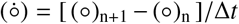 of the evolution of all five populations (o) = *S*, *E*, *I*_s_, *I*_a_, *R*. Here Δ*t* = *t*_n+1_ − *t*_n_ denotes the discrete time increment, usually Δ*t* = 1 day, and (o)_n+1_ and (o)_n_ denote the populations of the new and previous time steps. We apply an explicit time integration scheme to obtain the following discrete system of equations,

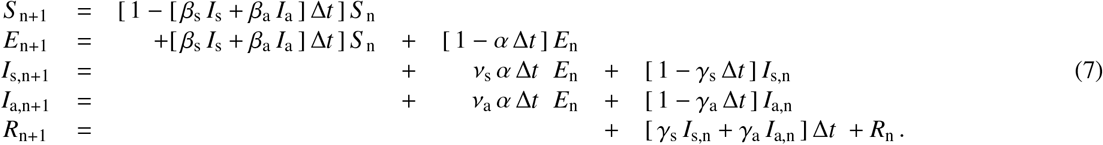

At each location, we begin our simulation on day *t*_0_, the day of the beginning of the local lockdown, with initial conditions, *S*_0_, *E*_0_, *I*_s0_, *I*_a0_, *R*_0_. We further assume that *S*_0_ = 1 − *E*_0_ − *I*_s0_ − *I*_a0_, *E*_0_ = [*A/C*_s_] [*I*_s0_ + *I*_a0_] [13], and *_ν_*_s_ *I*_a0_ = [1 − *v*_s_] *I*_s0_ to express all five initial conditions in terms of the initial symptomatic infectious population *I*_s0_, such that *S*_0_ = 1 − [1 + *A/C*_s_]/*v*_s_ *I*_s0_, *E*_0_ = [*A/C*_s_]/*v*_s_ *I*_s0_, *I*_a0_ = [1 − *v*_s_]/*v*_s_ *I*_s0_, and *R*_0_ = 0. From the solution of the time discrete SEIIR system (7), for each point in time, we calculate the of detected population,

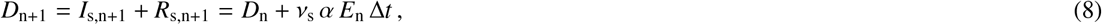

as the discrete sum of the symptomatic infectious and recovered populations *I*_s_,_n+1_ and *R*_s_,_n+1_ with 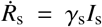, or, equivalently, as the explicit update of the previous detected population *D*_n_ with the new symptomatic infectious population, *v*_s_ *α E*_n_, that transitions from the exposed to the symptomatic infectious group. We compare the daily simulated detected population *D*(*t*) against the daily reported detected population 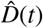, which we obtain by scaling the daily number of detected cases by the total population *N* for each location [5, 11, 27, 48, 49, 73].

### 2.2. Seroprevalence studies

We consider seroprevalence studies up to mid June 2020 where the representation ratio, the ratio between the amount of seroprevalence samples and the representative population, is larger than 0.02%. Subsequently, we draw the daily number of confirmed reported cases for Heinsberg, Ada County, New York City, Santa Clara County, Denmark, Geneva Canton, Netherlands, Rio Grande do Sul, and Belgium from online data repositories [5, 11, 27, 48, 49, 73]. The symptomatic fraction *v*_s_ follows from the ratio between the reported confirmed cases for the studied region and the amount of seroprevalence-estimated cases on the last day of the seroprevalence study. Table 1 summarizes the study date, number of samples, representation ratio, population, and symptomatic fraction of each location.

**Table 1:**
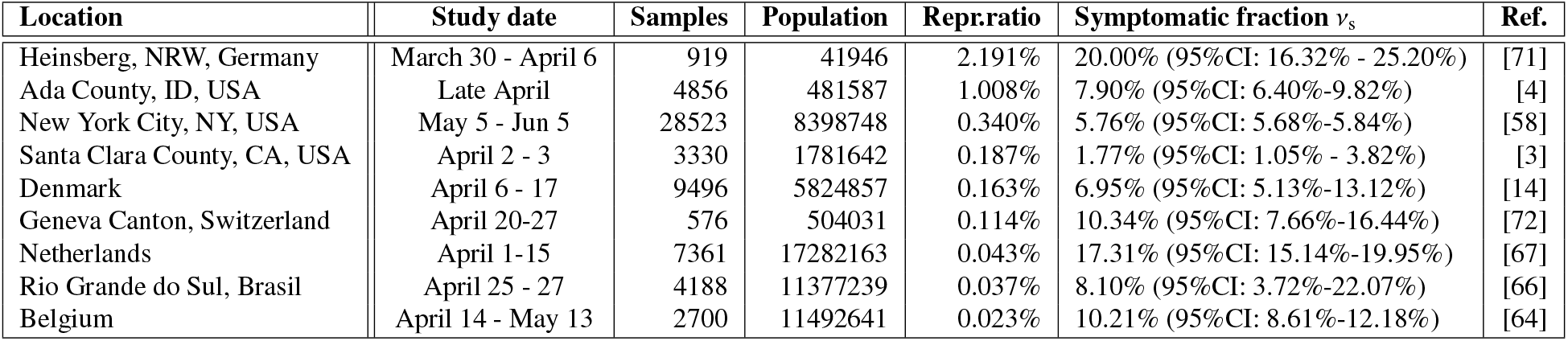
Seroprevalence studies with a representative population larger than 0.02%, location, study date, number of samples, representation ratio, population, and symptomatic fraction v_s_.

| Location | Study date | Samples | Population | Repr.ratio | Symptomatic fraction $\nu_s$ | Ref. |
| --- | --- | --- | --- | --- | --- | --- |
| Heinsberg, NRW, Germany | March 30 - April 6 | 919 | 41946 | 2.191% | 20.00% (95%CI: 16.32% - 25.20%) | [71] |
| Ada County, ID, USA | Late April | 4856 | 481587 | 1.008% | 7.90% (95%CI: 6.40%-9.82%) | [4] |
| New York City, NY, USA | May 5 - Jun 5 | 28523 | 8398748 | 0.340% | 5.76% (95%CI: 5.68%-5.84%) | [58] |
| Santa Clara County, CA, USA | April 2 - 3 | 3330 | 1781642 | 0.187% | 1.77% (95%CI: 1.05% - 3.82%) | [3] |
| Denmark | April 6 - 17 | 9496 | 5824857 | 0.163% | 6.95% (95%CI: 5.13%-13.12%) | [14] |
| Geneva Canton, Switzerland | April 20-27 | 576 | 504031 | 0.114% | 10.34% (95%CI: 7.66%-16.44%) | [72] |
| Netherlands | April 1-15 | 7361 | 17282163 | 0.043% | 17.31% (95%CI: 15.14%-19.95%) | [67] |
| Rio Grande do Sul, Brasil | April 25 - 27 | 4188 | 11377239 | 0.037% | 8.10% (95%CI: 3.72%-22.07%) | [66] |
| Belgium | April 14 - May 13 | 2700 | 11492641 | 0.023% | 10.21% (95%CI: 8.61%-12.18%) | [64] |

### 2.3. Bayesian learning

To calibrate our SEIIR model (1), we compare the model output, the simulated detected population *D*(*t*) for a given parameter set 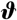, against the data, the reported detected population 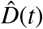 for each location [5, 11, 27, 48, 49, 73]. Our time-discrete SEIIR model (7) uses the following set of parameters,

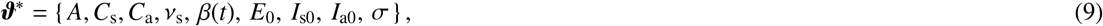

the latency period *A*, the symptomatic and asymptomatic infectious periods *C*_s_ and *C*_a_, the symptomatic fraction *v*_s_, the dynamic contact rate *β*(*t*), the initial exposed and infectious populations *E*_0_, *I*_s0_, and *I*_a0_, and the likelihood width o. To reduce the set of unknowns, we fix the latency period to *A* = 2.5 days and the symptomatic infectious period to *C*_s_ = 6.5 days [26, 37, 39, 62]. Since the asymptomatic infectious period *C*_a_ is unreported, we study three cases with *C*_a_ = 0.5,1.0,2.0 *C*_s_ resulting in infectious periods of *C*_a_ = 3.25, 6.5, and 13.0 days. We further express the initial exposed and asymptomatic infectious populations in terms of the symptomatic infectious population as *E*_0_ = [*A*/*C*_s_]/*v*_s_ *I*_s0_ [13] and *I*_a0_ = [1 − *v*_s_]/*v*_s_ *I*_s0_. This results in the following reduced set of parameters,

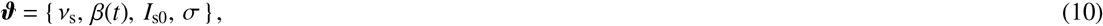

including the symptomatic fraction *v*_s_, the dynamic contact rate *β*(*t*), the initial symptomatic infectious population *I*_s0_, and the likelihood witdth *σ*.

#### Bayesian inference

We use Bayesian inference to estimate the set of parameters 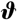 so that the statistics of the output of the model *D*(*t*) agree with that of the data 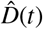 [41, 51]. We relate the prior, likelihood, and posterior using Bayes’ theorem [52],

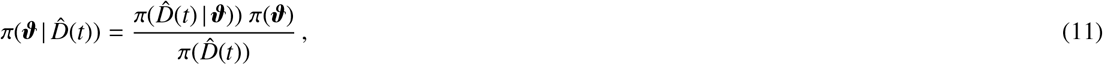

where 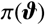 is the prior, i.e., the probability distribution of the model parameters, 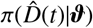 is the likelihood, i.e., the conditional probability of the data when the parameter set is fixed, 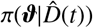 is the posterior, i.e., the conditional probability of the parameters for given data 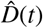, and 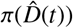 is the evidence.

#### Priors

For the prior probability distributions 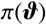, we assume weakly informative priors for our model parameters, the symptomatic fraction *v*_s_, the dynamic contact rate *β*(*t*), the initial symptomatic infectious population *I*_s0_, and the likelihood witdth *σ*. For the symptomatic fraction *v*_s_, we adopt the normal distributions from the individual local antibody seroprevalence studies of all nine locations [3,4, 14, 58, 64, 66, 67, 71, 72]. Specifically, we set 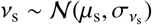, where *μ*_s_ and *σ*_s_ are the mean and standard deviation of the symptomatic fraction based on the ratio of the reported and seroprevalence-estimated numbers of confirmed cases for each location in Table 1. For the dynamic contact rate *β*(*t*), we postulate that its logarithm follows a Gaussian random walk, 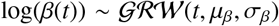. Specifically, this Gaussian random walk assumes a sequence of inter-dependent Gaussian distributions where the contact rate *β*(*t*_n+1_) on day *t*_n+1_ depends the contact rate *β*(*t*_n_) of the previous day *t*_n_ with the initial condition, 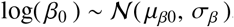, and the daily update, 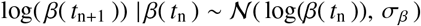, where *μ_β_*_0_ is the mean initial contact rate, *σ_β_* is the daily step width, and *μ_β_* is the overall drift between the initial value log(*β*(*t*_0_)) and the final value log(*β*(*t*_n_*_+_*_1_)). For the initial symptomatic infectious population *I*_s0_, we postulate a log-normal distribution, 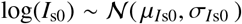, where we select a weakly informative prior for the mean 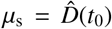, as the reported detected population on day *t*_0_ and a standard deviation of *σ*_s_ *=* 1. Table 2 summarizes our prior distributions and the SEIIR model parameters.

**Table 2:** Prior distributions and SEIIR model parameters.

| Parameter | Interpretation | Distribution |
| --- | --- | --- |
| $\nu_s$ | symptomatic fraction | see Table 1 [3, 4, 14, 58, 64, 66, 67, 71, 72] |
| $A$ | latent period | fixed ($A = 2.5$ days) [37, 39, 57] |
| $C_s$ | symptomatic infectious period | fixed ($C_s = 6.5$ days) [26, 39, 62] |
| $C_a$ | asymptomatic infectious period | fixed ($C_a = 3.25, 6.5, 13.0$ days) [26, 39, 62] |
| $\log(\beta(t))$ | dynamic contact rate | Gaussian random walk ($\mu = \mu_\beta, \sigma = \sigma_\beta$) |
| $\mu_{\beta 0}$ | initial contact rate | normal ($\mu = 0.0, \sigma = 1.0$) |
| $\mu_\beta$ | overall drift | normal ($\mu = 0.0, \sigma = 1.0$) |
| $\sigma_\beta$ | daily step width | half-normal ($\sigma = 0.02$) |
| $E_0$ | initial exposed | deterministic ($[A/C_s][I_{s0} + I_{a0}]$) [13] |
| $\log(I_{s0})$ | initial symptomatic | normal ($\mu = \hat{D}(t_0), \sigma = 1.0$) |
| $I_{a0}$ | initial asymptomatic | deterministic ($[1 - \nu_s]/\nu_s I_{s0}$) |
| $\sigma$ | likelihood width | halfCauchy ($\beta = 1.0$) |

#### Likelihood

For the likelihood 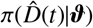, we introduce a likelihood function 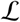 that evaluates the proximity between the model output *D*(*t*), the simulated detected population for a given parameter set 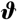, and the data 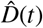, the relative reported detected population in probabilistic terms,

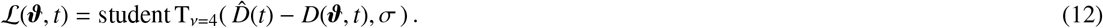

We choose a Student’s t-distribution because it resembles a Gaussian distribution around the mean with heavy tails, which make the method more robust with respect to outliers and reporting noise [8, 36]. Here *σ* represents the likelihood width for which we postulate a halfCauchy distribution, see Table 2. The likelihood,

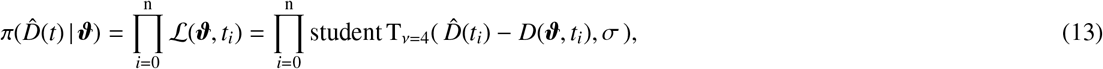

is simply the product of all likelihood functions 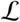 evaluated at each time *t_i_* for *i* = 0,1,…, n.

#### Posteriors

With the prior 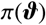 and the likelihood 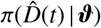, we estimate the posterior 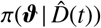 using Bayes’ theorem (11) [31]. Since we cannot describe the posterior distribution over the model parameters 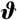 analytically, we adopt approximate-inference techniques to calibrate our model on the available data. We use the NO-U-Turn sampler (NUTS) [29], which is a type of Hamiltonian Monte Carlo algorithm as implemented in PyMC3 [61]. We use four chains. The first 4 times 500 samples serve to tune the sampler and are later discarded. The subsequent 4 times 1000 samples define the posterior distribution of the parameters 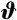. From the converged posterior distribution 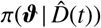, we sample multiple combinations of parameters 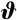 that describe the time evolution of detected population 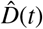. Using these posterior samples, we quantify the uncertainty of each parameter. As such, each parameter set 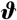 provides a set of values for the symptomatic fraction *v*_s_, the initial symptomatic populations *I*_s0_, and the dynamic contact rate *β*(*t*). From these values, we quantify the effective reproductive number *R*(*t*) using equation (5) and the time evolution of the susceptible, exposed, symptomatic infectious, asymptomatic infectious and recovered populations, *S*(*t*), *E*(*t*), *I*_s_(*t*), *I*_a_(*t*), and *R*(*t*) using equations (7) and report their values with the associated 95% credible interval [50].

#### Hierarchical modeling

To estimate the asymptomatic infectious period *C*_a_, we create a hierarchical model and analyze the confirmed case data from all nine locations in Table 1. To prevent overfitting, we postulate that the initial reproduction number *R*_0_ is the same for all nine locations and remains constant during a two-week time window prior to lockdown. This implies that the contact rate *β* is static and constant. We assume a normal distribution for the basic reproduction number 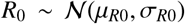 with mean *μ_R_*_0_ = 3.87 and standard deviation *σ_R_*_0_ *=* 0.44 [17]. This defines the initial contact rate *β*_0i_ at each location, i = 1,…, 9, as *β*_0i_ = *R*_0_ (*v*_si_/*C*_s_ + (1 − *v*_si_)/*C*_a_i). We create a hyper-distribution for the overall asymptomatic infectious period, 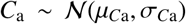, with mean *μ_C_*_a_ = 6.5 and standard deviation 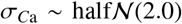, and assume that each local asymptomatic infectious period 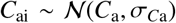 is drawn from this hyperdistribution. All other parameters are similar to the previous section. Table 3 summarizes the prior distributions and the SEIIR model parameters of our hierarchical model to infer the asymptomatic infectious period *C*_a_.

**Table 3:**
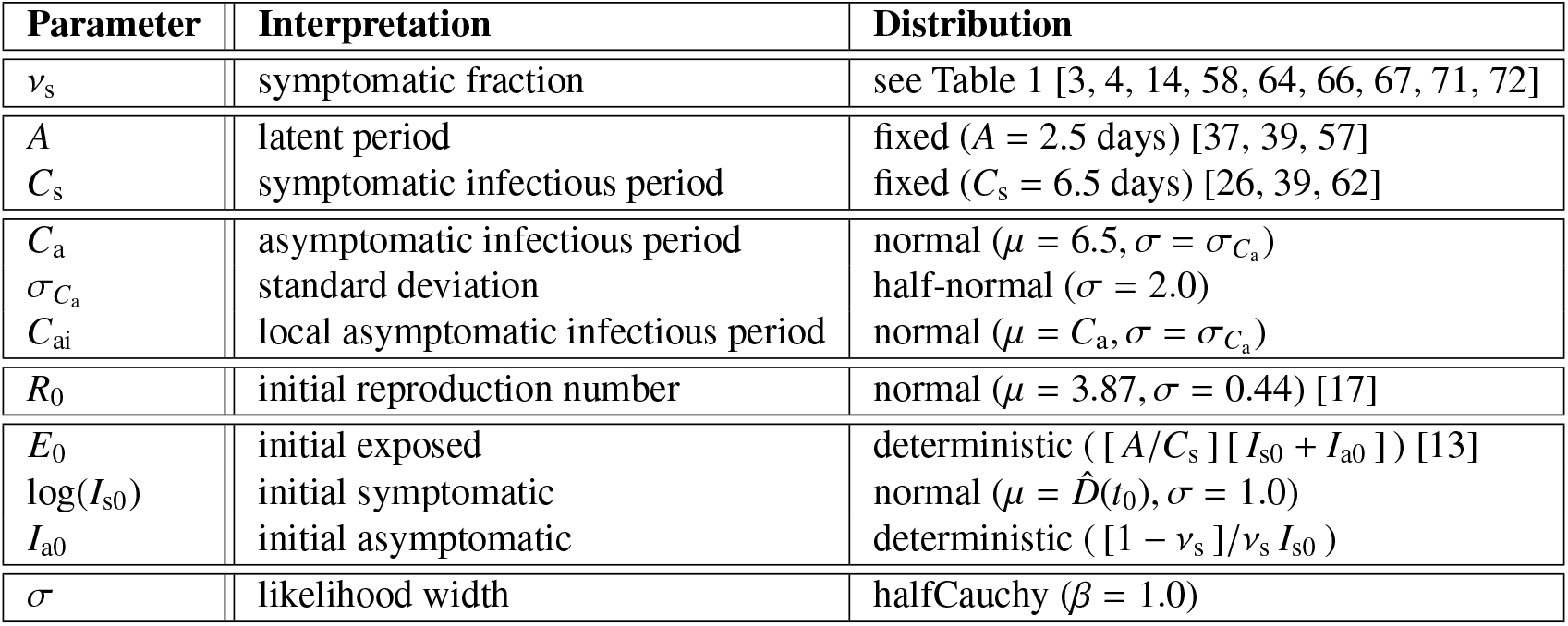
Prior distributions and SEIIR model parameters to hierarchically infer the asymptomatic infectious period *C*_a_.

To estimate the hierarchical asymptomatic infectious period *C*_a_, we use Bayesian inference. Following equation (11), we relate the prior, likelihood, and posterior, with the only difference that the simulated and reported detected populations *D*_i_(*t*) and 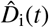 are now vectors that contain the detected populations of all *i =* 1,…, 9 locations. The likelihood 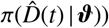 becomes the product of the all likelihood functions 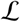 evaluated for a period of n = 14 days prior to lockdown,

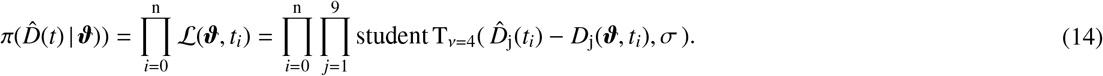

For each location, we estimate the posterior 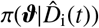 using the NO-U-Turn sampler (NUTS) [29] in PyMC3 [61]. From the converged posterior distribution, we sample multiple 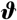 combinations to quantify the evolution of the reproduction number *R*_0_ and contact rate *β* at each location, and the associated susceptible, exposed, symptomatic and asymptomatic infectious, and recovered populations with their associated 95% credible intervals.

#### Outbreak date

For each sample from the posterior distribution, we use the infered initial exposed and asymptomatic infectious populations *E*_0_ and *I*_a0_ to estimate the date of the very first COVID-19 case in Santa Clara County [63]. Specifically, for each parameter set, we create an SEIIR model and assume that the outbreak begins with one single asymptomatic infectious individual. We fix the latency and symptomatic infectious periods to *A =* 2.5 days and *C*_s_ = 6.5 days and impose the hierarchically estimated infectious period *C*_a_ from the previous section. For each posterior sample for the exposed, symptomatic infectious, and asymptomatic infectious population size on the lockdown date, March 16, 2020, we use the Nelder-Mead optimization method [21] to find the most probable outbreak origin date. Specifically, we solve the SEIIR model forward in time using an explicit time integration, starting from various start dates before March 16, 2020, and iteratively minimize the difference between the computed exposed, symptomatic, and asymptomatic infectious populations and the sample’s actual exposed, symptomatic, and asymptomatic infectious populations. We concomitantly fit a static contact rate *β*, which is bounded between zero and the posterior sample’s estimated contact rate *β*_0_ at the beginning of the simulation *t*_0_. Repeating this process for each sample of the Bayesian inference generates a distribution of possible origin dates. From this distribution, we compute the most probable origin date and its uncertainty.

## 3. Results

### 3.1. Outbreak dynamics of COVID-19 in Santa Clara County

Figure 3 illustrates the outbreak dynamics of COVID-19 in Santa Clara County. To explore the effects of the asymptomatic infectious period *C*_a_ on the initial effective reproduction number, we start the simulations on March 6, 2020, 10 days before the local lockdown date. The black dots highlight the reported detected population 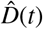 from this day forward. Based on these data points, we learn the posterior distributions of our SEIIR model parameters for fixed latent and symptomatic infectious periods *A* = 2.5 days and *C*_s_ = 6.5 days, and for three asymptomatic infectious periods, *C*_a_ = 3.25, 6.5, and 13.0 days, from left to right. The gray and green-purple regions highlights the 95% credible intervals on the confirmed cases *D*(*t*), top row, and the contact rate *β*(*t*), bottom row, based on the reported cases D(t), while taking into account uncertainties on the symptomatic infectious fraction *v*_s_, and the initial exposed and infectious populations *E*_0_, *I*_s0_, and *I*_a0_. The red, orange, and green histograms on the middle row display the inferred initial exposed and infectious populations, *E*_0_, *I*_s0_, and *I*_a0_, for the three different asymptomatic infectious periods. The graphs confirm that our dynamic SEIIR epidemiology model is capable of correctly capturing the gradual flattening and deflattening of the curve of confirmed cases in agreement with the decrease in new cases reported in Santa Clara County, top row. The consistent downward trend of the contact rate *β*(*t*) after the lockdown date (March 16, 2020) quantifies the efficiency of public health interventions. The different magnitudes in the contact rate highlight the effect of the three different asymptomatic infectious periods *C*_a_: For larger asymptomatic infectious periods *C*_a_, from left to right, to explain the same number of confirmed cases *D*(*t*) *= I*_s_(*t*) + *R*_s_(*t*), the contact rate *β*(*t*) has to decrease. On March 6, 2020, the mean contact rate *β*(*t*) was 0.688 (95% CI: 0.612 - 0.764) for an infectious period of *C*_a_=3.25 days, 0.529 (95% CI: 0.464 - 0.593) for *C*_a_= 6.5 days, and 0.463 (95% CI: 0.403 - 0.523) for *C*_a_ =13.0 days. By March 16, 2020, the day Santa Clara County announced the first county-wide shelter-in-place order in the entire United States, these contact rates *β*(*t*) were 0.491 (95% CI: 0.462 - 0.523) for an infectious period of *C*_a_=3.25 days, 0.328 (95% CI: 0.305 - 0.352) for *C*_a_= 6.5 days, and 0.252 (95% CI: 0.234 - 0.271) for *C*_a_=13.0 days.

**Figure 3:**
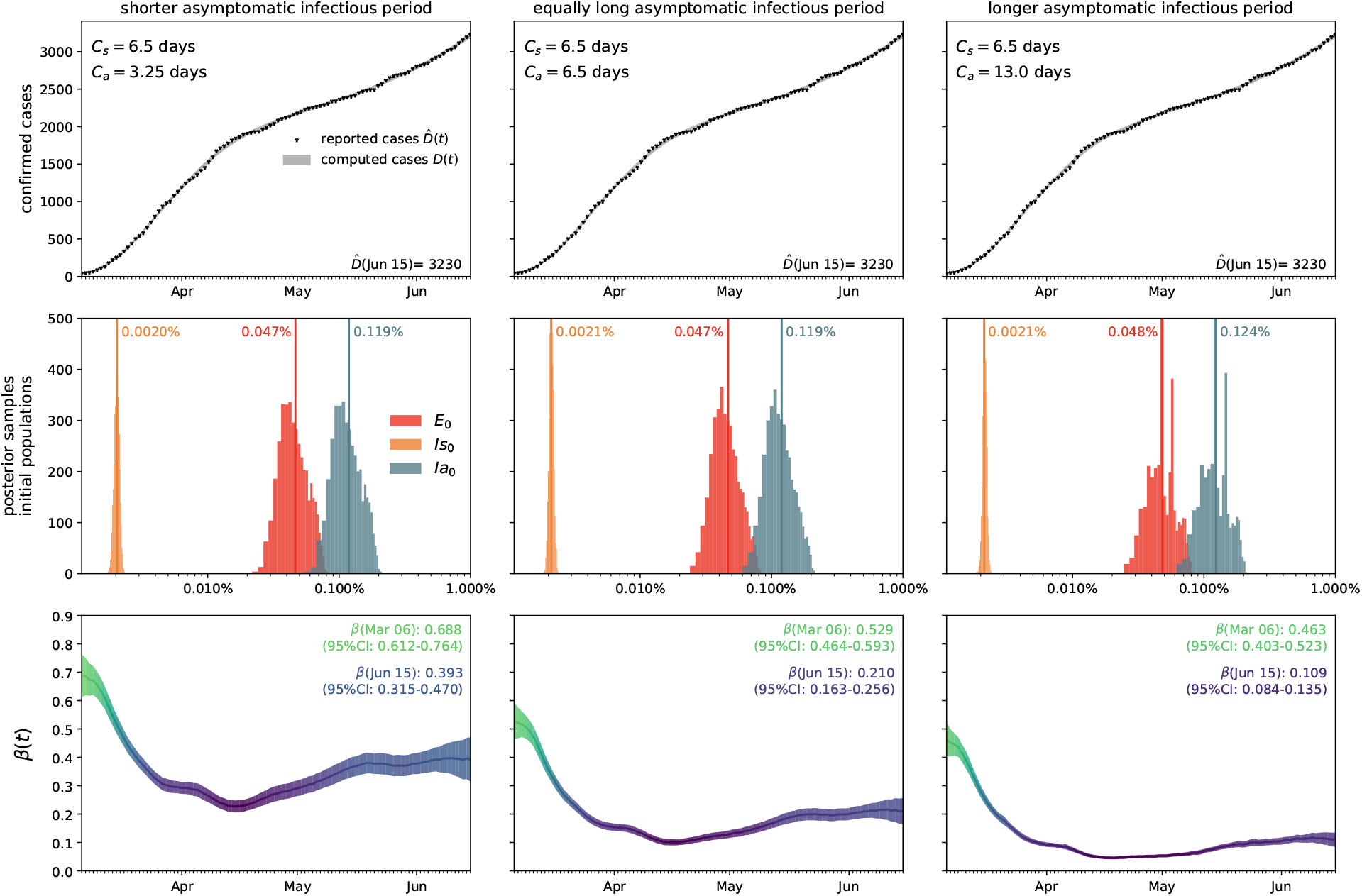
Outbreak dynamics of COVID-19 in Santa Clara County. The simulation learns the time-varying contact rate *β*(*t*) for fixed latent and symptomatic infectious periods *A* = 2.5 days and *C*_s_ = 6.5 days, and for three asymptomatic infectious periods *C*_a_ = 3.25 days, 6.5 days, and 13.0 days (from left to right). Computed and reported confirmed cases in Santa Clara County, *D*(*t*) = *I*_s_(*t*) + *R*_s_(*t*) and 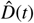 (top), initial exposed and infectious populations, *E*_0_, *I*_s0_, and *I*_a0_ (middle), and dynamic contact rate, *β*(*t*) (bottom). The gray and green-blue regions highlight the 95% credible intervals on the confirmed cases *D*(*t*) (top) and the contact rate *β*(*t*) (bottom) based on the reported cases 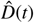, while taking into account uncertainties on the fraction of the symptomatic infectious population *v*_s_ = *I*_s_ /*I*, and the initial exposed and infectious populations *E*_0_, *I*_s0_, and *I*_a0_.

### 3.2. Effect of asymptomatic transmission ofCOVID-19 in Santa Clara County

Figure 4 visualizes the effect of asymptomatic transmission in Santa Clara County. The simulation learns the dynamic contact rate *β*(*t*), and with it the time-varying effective reproduction number *R*(*t*), top row, for three asymptomatic infectious periods *C*_a_ = 3.25 days, 6.5 days, and 13.0 days, from left to right. The effective reproduction number *R*(*t*) follows a similar downward trend as the contact rate *β*(*t*). For larger asymptomatic infectious periods *C*_a_, from left to right, since *R*(t) *= C*_s_*β*(*t*)*/*[*v*_s_ + *v*_a_ *C*_s_/*C*_a_], as *C*_a_ increases, *C*_s_/*C*_a_ decreases, and *R*(*t*) increases. Since *R*(*t*) represents the number of new infections from a single case, a decrease below *R*(*t*) < 1 implies that a single infectious individual infects less than one new individual, which indicates that the outbreak decays. The dashed vertical lines indicate the date *R*(*t*) = 1 during which one infectious individual, either symptomatic or asymptomatic, infects on average one other individual. For an asymptomatic infectious period of *C*_a_=3.25 days, it took until March 28 before Santa Clara County managed to get *R*(*t*) below 1 for the first time after the outbreak. For *C*_a_=6.5 days, this only occurred by April 1 and for *C*_a_=13.0 days, this occurred on April 8, 2020. This confirms our intuition that, the larger the asymptomatic infectious period *C*_a_, for example because asymptomatic individuals will not isolate as strictly as symptomatic individuals, the higher the effective reproduction number *R*(*t*), and the more difficult it will be to control *R*(*t*) by public health interventions. For each of the three cases, the symptomatic infectious, asymptomatic infectious, and recovered population, are shown in the bottom row. For larger asymptomatic infectious periods *C*_a_, from left to right, the total infectious population *I* increases and its maximum occurs later in time. Specifically, the maximum infectious population since March 6, 2020 amounts to 0.70% (95% CI: 0.43%-0.97%) on March 28, 2020 for *C*_a_ = 3.25 days, 1.23% (95% CI: 0.72%-1.75%) on April 2, 2020 for *C*_a_ = 6.5 days, and 2.10% (95% CI: 1.25%-2.94%) on April 7, 2020 for *C*_a_ = 13.0 days. For larger asymptomatic infectious periods *C*_a_, from left to right, the recovered population *R* decreases. Specifically, on June 15, 2020, the recovered population *R* amounts to 10.85% (95% CI: 7.06%-16.09%) for an infectious period of *C*_a_ =3.25 days, 10.20% (95% CI: 6.07%-14.90%) for *C*_a_ =6.5 days, and 9.90% (95% CI: 6.07%-13.93%) for *C*_a_ =13.0 days. Similarly, and important when considering different exit strategies, the total infectious population, *I* = *I*_s_ + *I*_a_, on June 15, 2020 is estimated to 0.39% (95% CI: 0.24%-0.58%) for *C*_a_ =3.25 days, 0.68% (95% CI: 0.40%-1.01%) for *C*_a_ =6.5 days, and 1.25% (95% CI: 0.73%-1.77%) for *C*_a_ =13.0 days.

**Figure 4:**
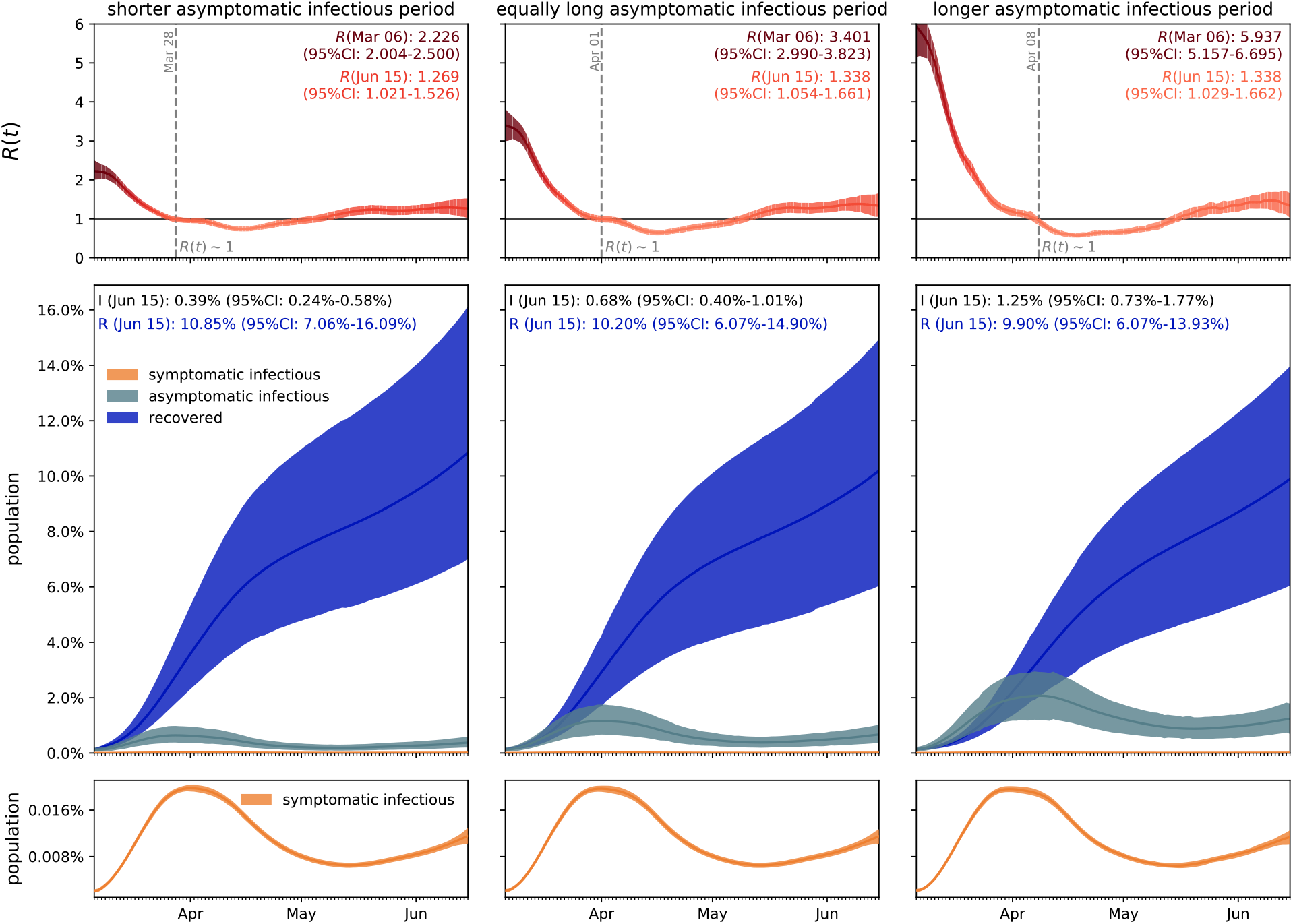
Effect of asymptomatic transmission of COVID-19 in Santa Clara County. The simulation learns the time-varying contact rate *β*(*t*), and with it the time-varying effective reproduction number *R*(*t*), for fixed latent and symptomatic infectious periods *A =* 2.5 days and *C*_s_ = 6.5 days, and for three asymptomatic infectious periods *C*_a_ = 3.25 days, 6.5 days, and 13.0 days (from left to right). The downward trend of the effective reproduction number *R*(*t*) reflects the efficiency of public health interventions (top row). The dashed vertical lines mark the critical time period during which the effective reproductive reproduction number fluctuates around *R*(*t*) *=* 1. The simulation predicts the symptomatic infectious, asymptomatic infectious, and recovered populations *I*_s_, *I*_a_, and *R* (bottom row). The colored regions highlight the 95% credible interval for uncertainties in the number of confirmed cases D, the fraction of the symptomatic infectious population *v*_s_ = *I*_s_*/I*, the initial exposed population *E*_0_ and the initial infectious populations *I*_s0_ and *I*_a0_.

### 3.3. Outbreak dynamics of COVID-19 worldwide

Figure 5 illustrates the hierarchical asymptomatic infectious period *C*_a_ estimation for nine locations that reported COVID-19 antibody prevalence in a representative sample of the population: Heinsberg (NRW, Germany), Ada County (ID, USA), New York City (NY, USA), Santa Clara County (CA, USA), Denmark, Geneva Canton (Switzerland), Netherlands, Rio Grande do Sul (Brazil) and Belgium. This estimate assumes an initial fixed reproduction number *R*_0_ = 3.87 (95%CI: 3.01-4.66) during a two-week window before lockdown which results in an asymptomatic infectious period of *C*_a_ = 5.76 (95%CI: 3.59-8.09) days. With this value, we simulate the outbreak dynamics of COVID-19 in all nine locations. Figure 6 illustrates the learnt effective reproduction number *R*(*t*), and the symptomatic and asymptomatic infectious populations *I*_s_ and *I*_a_, and the recovered population *R* for all nine locations. Here, we assume fixed latent and infectious periods of *A* = 2.5 days, *C*_s_ = 6.5 days, and the hierarchical asymptomatic infectious period *C*_a_ = 5.76 (95%CI: 3.59-8.09) days from Figure 5.

**Figure 5:**
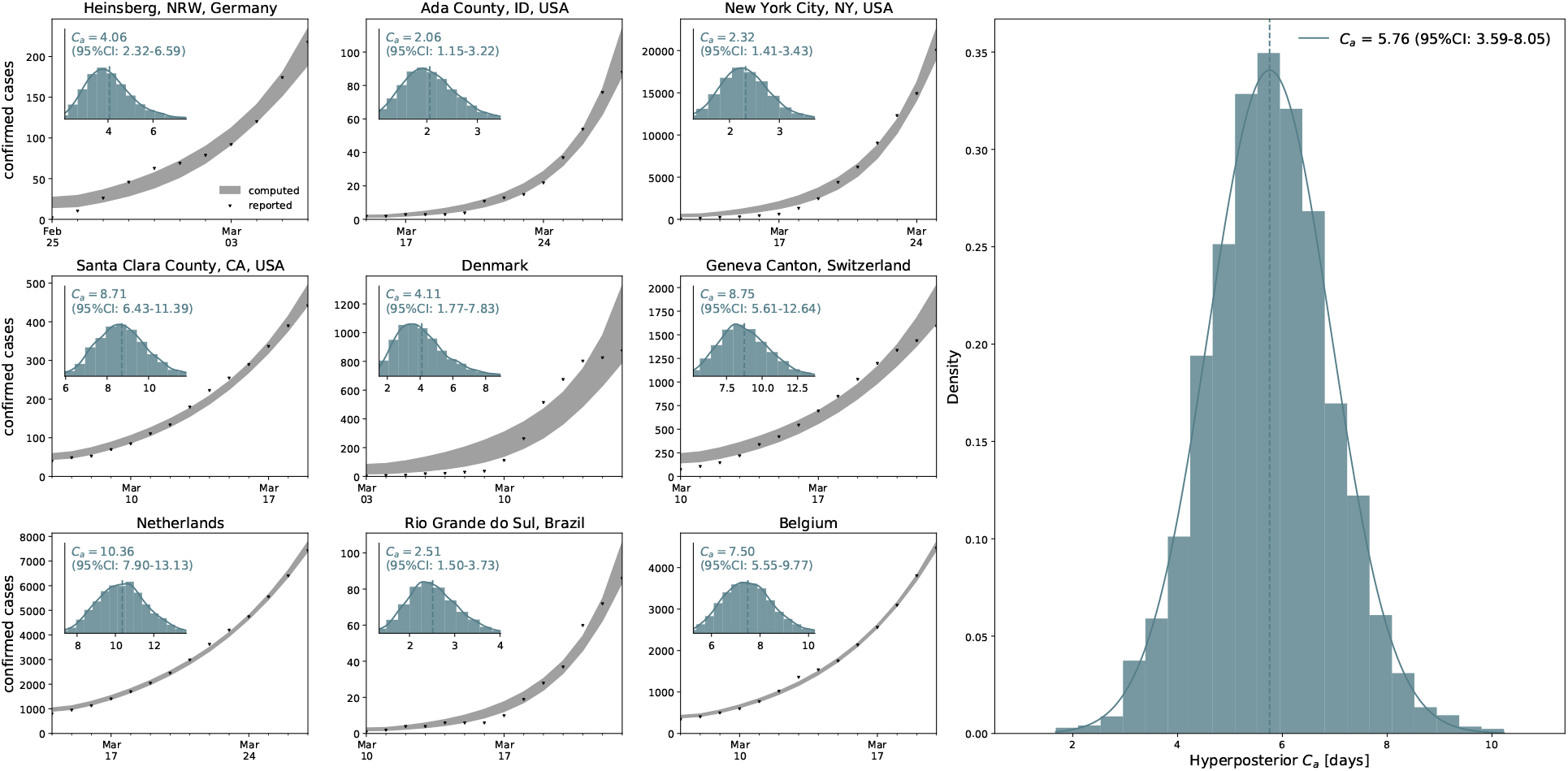
Hierarchical asymptomatic infectious period *C*_a_ estimation. Assuming an initial fixed reproduction number *R*_0_ = 3.87 (95%CI: 3.01-4.66) [17], the simulation generates histograms of the asymptomatic infectious period *C*_a_ for each location based on the location-specific symptomatic fraction *v*_s_. The black dots and grey regions represent the reported and simulated detected cases 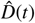 and *D*(*t*) respectively. The hierarchical hyperdistribution for the asymptomatic infectious period results in *C*_a_ = 5.76 (95%CI: 3.59-8.09) days, right histogram.

For all nine locations, the calculated metrics display the fine balance between the dynamics of the effective reproduction number and the control of the epidemic outbreak. The downward trend of the effective reproductive number *R*(*t*) quantifies how fast each location managed to control the spreading of COVID-19. The dashed vertical line indicates the first time each location managed to lower the effective reproduction below *R*(*t*) = 1 after lockdown. This critical transition occurred on March 20 for Heinsberg, April 4 for Ada County, April 28 for New York City, March 30 for Santa Clara County, April 8 for Denmark, April 2 for Geneva, April 14 for the Netherlands, June 8 for Rio Grande do Sul and April 11 for Belgium. Based on our simulations, the maximum infectious population size amounted to 3.54% (95% CI: 2.96%-4.11%) on March 19 in Heinsberg, 0.47% (95% CI: 0.41%-0.55%) on May 5 in Ada County, 6.11% (95% CI: 5.99%-6.21%) on April 13 in New York City, 1.11% (95% CI: 0.69%-1.65%) on March 29 in Santa Clara County, 0.38% (95% CI: 0.27%-0.50%) on April 9 in Denmark, 2.10% (95% CI: 1.65%-2.69%) on April 2 in Geneva, 0.62% (95% CI: 0.56%-0.68%) on April 15 in the Netherlands, 0.28% (95% CI: 0.14%-0.48%) on June 8 in Rio Grande do Sul, and 0.70% (95% CI: 0.62%-0.78%) on April 11 in Belgium. On Jun 15, 2020, the estimated recovered population reached 24.15% (95% CI: 20.48%-28.14%) in Heinsberg, NRW, Germany 2.40% (95% CI: 2.09%-2.76%) in Ada County, ID, USA 46.19% (95% CI: 45.81%-46.60%) in New York City, NY, USA 11.26% (95% CI: 7.21%-16.03%) in Santa Clara County, CA, USA 3.09% (95% CI: 2.27%-4.03%) in Denmark 12.35% (95% CI: 10.03%-15.18%) in Geneva Canton, Switzerland 5.24% (95% CI: 4.84%-5.70%) in Netherlands 1.53% (95% CI: 0.76%-2.62%) in Rio Grande do Sul, Brazil 5.32% (95% CI: 4.77%-5.93%) in Belgium.

### 3.4. Estimating the outbreak date

Figure 7 shows the estimated outbreak date of COVID-19 in Santa Clara County. For fixed latent and symptomatic infectious periods *A* = 2.5 days and *C*_s_ = 6.5 days, and for the hierarchically estimated asymptomatic infectious periods *C*_a_ = 5.76 (95%CI: 3.59-8.09) days, the graph highlights the estimated date of the first COVID-19 case in the county. Based on the reported case data from March 16, 2020 onward, and taking into account uncertainty on the fraction of the symptomatic infectious population *v*_s_, on the initial exposed population *E*_0_, and on the initial symptomatic and asymptomatic infectious populations *I*_s0_ and *I*_a0_, we systematically backtracked the date of the first undetected infectious individual. Our results suggest that the first case of COVID-19 in Santa Clara County dates back to January 20, 2020 (95% CI: December 29, 2019 - February 13, 2020).

## 4. Discussion

A key question in understanding the outbreak dynamics of COVID-19 is the dimension of the asymptomatic population and its role in disease transmission. Throughout the past months, dozens of studies have been initiated to quantify the fraction of the general population that displays antibody prevalence but did not report symptoms of COVID-19. Here we assume that this subgroup of the population has been infected with the novel coronavirus, but has remained asymptomatic, or only displayed mild symptoms that were not directly reported in the context of COVID-19. We collectively map this subgroup into an asymptomatic population and additively decompose the total infectious population, *I* = *I*_s_ + *I*_a_, into a symptomatic group *I*_s_ and an asymptomatic group *I*_a_. We parameterize this decomposition in terms of a single scalar valued parameter, the symptomatic fraction *v*_s_. Within this paradigm, we can conceptually distinguish two scenarios: the special case for which both subgroups display identical contact rates *β* latent periods *A*, and infectious periods *C*, and the general case for which these transition dynamics are different.

### For comparable dynamics, the size of the asymptomatic population does not affect overall outbreak dynamics

For the special case in which both subgroups display identical contact rates *β*, latent rates *α*, and infectious rates *γ* [79], our study shows that the overall outbreak dynamics can be represented by the classical SEIR model [28] using equations (6). Importantly, however, since the reported case data only reflect the symptomatic infectious and recovered groups *I*_s_ and *R*_s_, the true infectious and recovered populations *I* = *I*_s_/*v*_s_ and *R = R*_s_/*v*_s_ could be about an order of magnitude larger than the SEIR model predictions. From an individual’s perspective, a smaller symptomatic group *v*_s_, or equivalently, a larger asymptomatic group *v*_a_ = [1 − *v*_s_], could have a personal effect on the likelihood of being unknowingly exposed to the virus, especially for high-risk populations: A larger asymptomatic fraction *v*_a_ would translate into an increased risk of community transmisson and would complicate outbreak control [16]. From a health care perspective, however, the special case with comparable transition dynamics would not pose a threat to the health care system since the overall outbreak dynamics would remain unchanged, independent of the fraction *v*_a_ of the asymptomatic population: A larger asymptomatic fraction would simply imply that a larger fraction of the population has already been exposed to the virus-without experiencing significant symptoms-and that the true hospitalization and mortality rates would be much lower than the reported rates [30].

**Figure 6:**
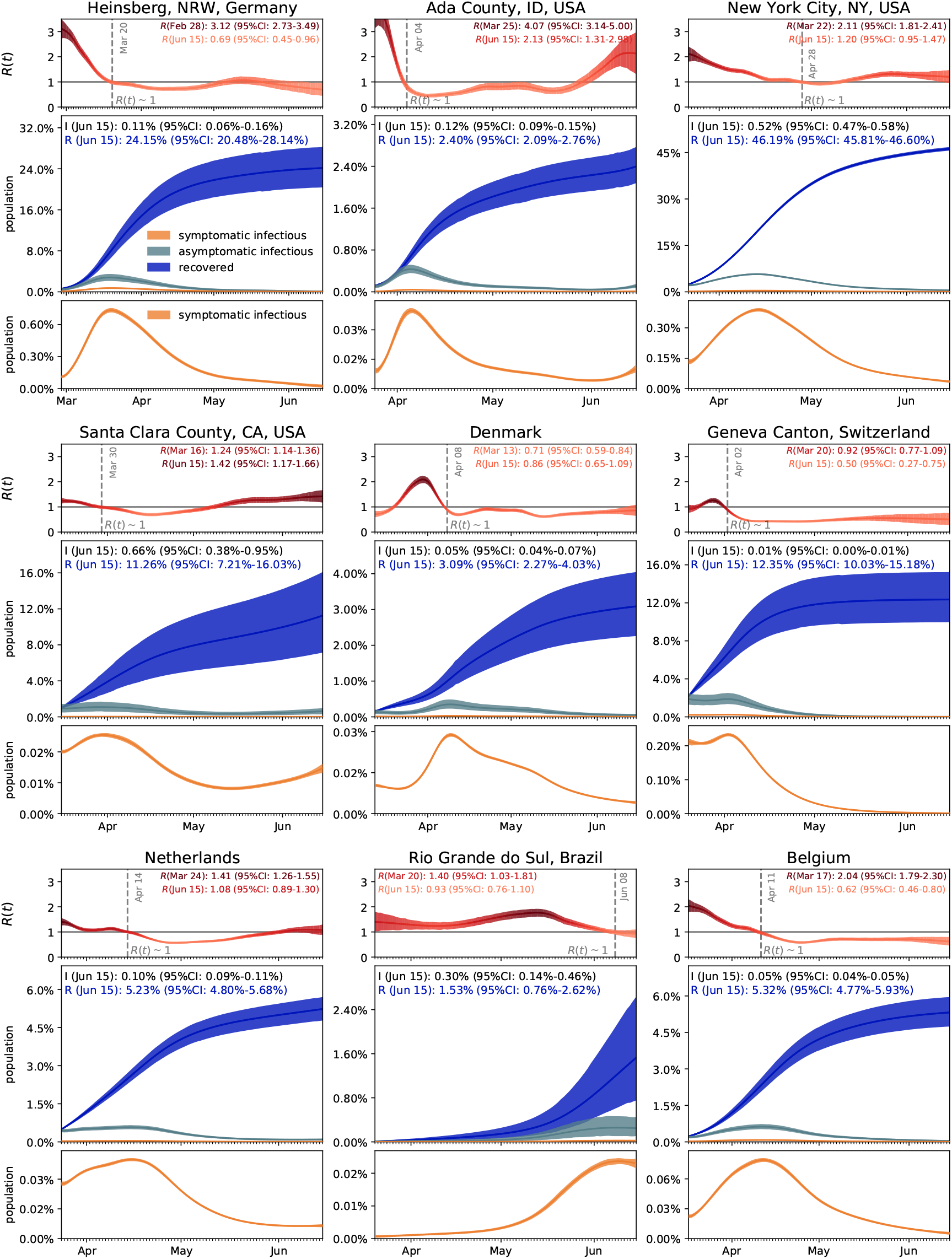
Outbreak dynamics of COVID-19 worldwide. Dynamic effective reproduction number R(t) and symptomatic, asymptomatic, and recovered populations at all nine locations. The simulation learns the time-varying contact rate *β*(*t*), and with it the time-varying effective reproduction number *R*(*t*), to predict the symptomatic infectious, asymptomatic infectious, and recovered populations *I*_s_, *I*_a_, and *R*, for fixed latent and infectious periods *A* = 2.5 days, *C*_s_ = 6.5 days, the hierarchical asymptomatic infectious period *C*_a_ = 5.76 (95%CI: 3.59-8.09) days from Figure 5. The dashed vertical lines mark the the first time each location managed to lower the effective reproduction below *R*(*t*) = 1 after lockdown. The colored regions highlight the 95% credible interval for the effective reproductive number *R*(*t*) (top), the symptomatic and asymptomatic populations *I*_s_ and *I*_a_, and the recovered population *R* (bottom plots), for uncertainties in the number of confirmed cases *D*, the fraction of the symptomatic infectious population *v*_s_, the initial exposed population *E*_0_, and the initial infectious populations *I*_s0_ and *I*_a0_.

**Figure 7:**
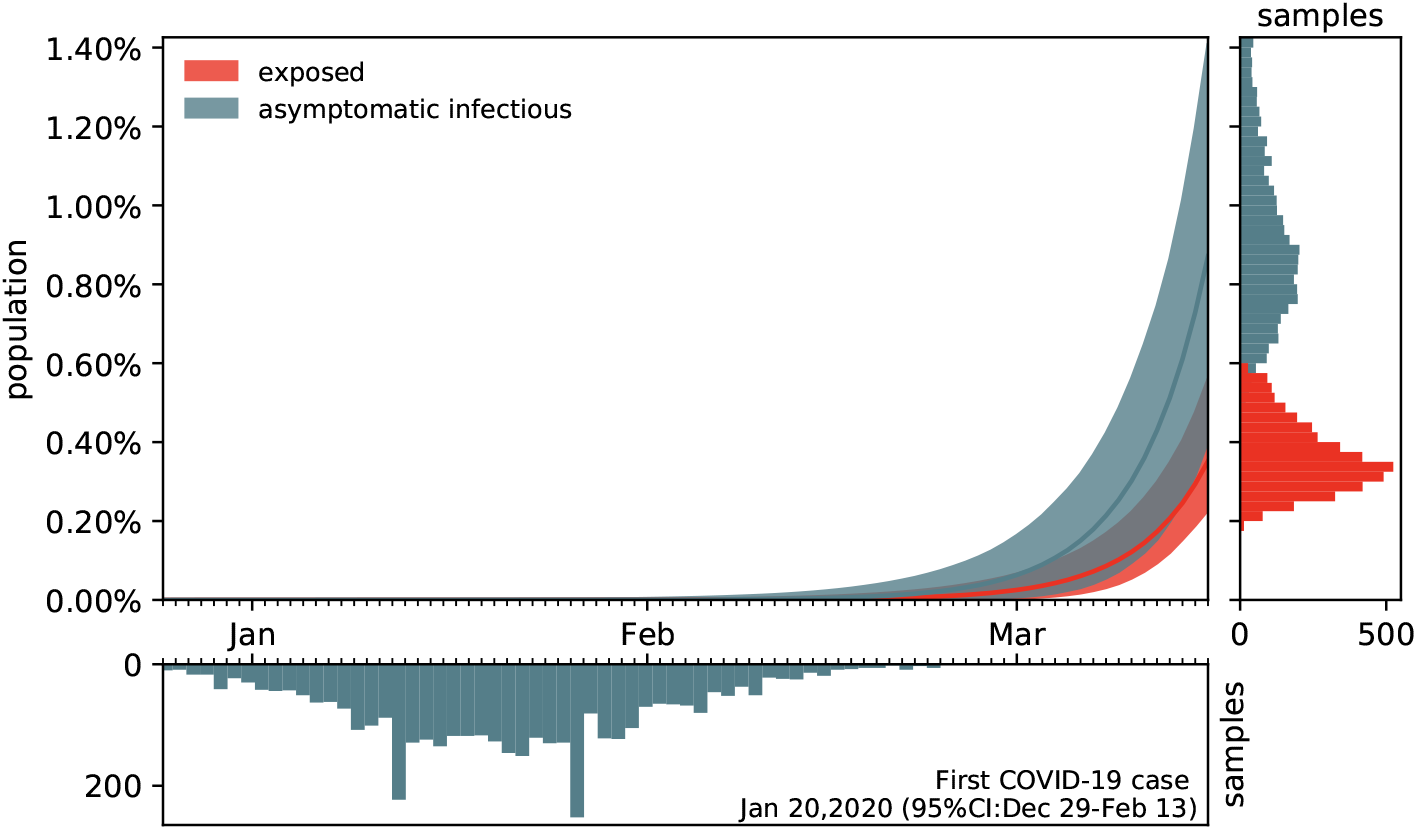
Estimating the outbreak date of COVID-19 in Santa Clara County varying asymptomatic infectious periods *C*_a_. Estimated date of the first COVID-19 case in Santa Clara County for fixed latent and symptomatic infectious periods *A =* 2.5 days and *C*_s_ = 6.5 days, and for the hierarchical asymptomatic infectious period *C*_a_ = 5.76 (95%CI: 3.59-8.09) days from Figure 5. The colored regions in the main plot highlight the 95% credible interval for the time evolution of the exposed and asymptomatic infectious populations *E* and *I*_a_ estimated based on the reported cases 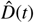 from March 16, 2020 onward and taking into account uncertainties on the fraction of the symptomatic infectious population *v*_s_ = *I*_s_/*I*, and the exposed and asymptomatic infectious populations *E*_0_ and *I*_a0_ on March 16, 2020 (right plot). The bottom plot histogram shows the distribution of the most probable origin dates to January 20, 2020 (95% CI: December 29, 2019 - February 13, 2020).

### For different dynamics, the overall outbreak dynamics depend on both size and infectiousness of the asymptomatic group

For the general case in which the transition rates for the symptomatic and asymptomatic groups are different, the overall outbreak dynamics of COVID-19 become more unpredictable, since little is known about the dynamics of the asymptomatic population [54]. To study the effects of different dynamics between the symptomatic and asymptomatic groups, we decided to collectively represent a lower infectivity of the asymptomatic population through a smaller infectious period *C*_a_ < *C*_s_ and a lack of early isolation of the asymptomatic population through a larger infectious period *C*_a_ > *C*_s_, while, for simplicity, keeping the latent period A and contact rate *β* similar across both groups [39]. Our study shows that the overall reproduction number, *R*(*t*) *=* [*C*_a_ *C*_s_]/[*v*_s_ *C*_a_ + *v*_a_ *C*_s_] *β*(*t*), and with it the outbreak dynamics, depend critically on the fractions of the symptomatic and asymptomatic populations *v*_s_ and *v*_a_ and on the ratio of the two infectious periods *C*_s_ and *C*_a_. To illustrate these effects, we report the results for three different scenarios where the asymptomatic group is half as infectious, *C*_a_ = 0.5 *C*_s_, equally infectious, *C*_a_ = *C*_s_, and twice as infectious *C*_a_ = 2.0 *C*_s_ as the symptomatic group for Santa Clara County. The second case, the middle column in Figures 3 and 4, corresponds to the special case with comparable dynamics and similar parameters. Our learnt asymptomatic infectious periods of *C*_a_ = 5.76 days in Figure 5 suggest that *C*_a_ is smaller than the symptomatic infectious period of *C*_s_ = 6.5 days and that the asymptomatic population is slightly less infectious as the symptomatic population. This can be a combined effect of being less contagious but, at the same time, having more contacts, since asymptomatic individuals, by definition, do not realize that they are spreading the disease [38].

### Dynamic contact rates are a metric for the efficiency of public health interventions

Classical SEIR epidemiology models with static parameters are well suited to model outbreak dynamics under unconstrained conditions and predict how the susceptible, exposed, infectious, and recovered populations converge freely toward the endemic equilibrium [28]. However, they cannot capture changes in disease dynamics and fail to converge towards a temporary equilibrium before the entire population has become sufficiently immune to prevent further spreading [57]. To address this limitation, we introduce a time-dependent contact rate *β*(*t*), which we learn dynamically from the reported case data. Figures 3 and 6 demonstrate that our approach can successfully identify a dynamic rate that not only decreases monotonically, but is also capable of reproducing local contact fluctuations [43]. With this dynamic contact rate, our model can capture the characteristic S-shaped COVID-19 case curve that plateaus before a large fraction of the population has been affected by the disease, resembling a Gompertz function. It also naturally captures a potential regrowth of the contact rate *β*(*t*), and with it the effects of a second wave [45]. Previous studies have inferred discrete date points at which the contact rates vary [8] or used sliding windows over the amount of novel reported infections [55] to motivate dynamic contact rates. As such, our framework provides a model-based method for statistical inference of virus transmissibility: It naturally learns the most probable contact rate from the changing time evolution of new confirmed cases and concomitantly quantifies the uncertainty on that estimation.

### The dynamics of the asymptomatic population affect the effective reproduction number

Our analysis in equations (4) and (5) and our simulations in Figure 4 illustrate how asymptomatic transmission affects the effective reproduction number, and with it the outbreak dynamics of COVID-19. Our results show that, the larger the infectious period *C*_a_ of the asymptomatic group, the larger the initial effective reproduction number, *R*(*t*) = *C*_s_*β*(*t*)/[*v*_s_ + *v*_a_ *C*_s_/*C*_a_], and the later the drop of *R*(*t*) below the critical value of one. A recent study analyzed the dynamics of the asymptomatic population in three consecutive windows of two weeks during the early outbreak in China [40]. The study found relatively constant latency and infectious periods *A* and *C*, similar to our assumption, and a decrease in the contact rate *β* = 1.12,0.52,0.35 days^−1^ and in the effective reproduction number *R*(*t*) = 2.38,1.34,0.98, which is consistent with our results. However, rather than assuming constant outbreak parameters within pre-defined time windows, our study learns the effective reproduction number dynamically, in real time, from the available data. Figures 4 and 6 demonstrate that we can successfully learn the critical time window until *R*(*t*) drops below 1, which, in Santa Clara County, took till March 28 for *C*_a_ =3.25 days, till April 1 for *C*_a_ = 6.5 days and till April 8 for *C*_a_ = 13.0 days. Our findings are consistent with the observation that the basic reproduction number will be over-estimated if the asymptomatic group has a shorter generation interval, and underestimated if it has a longer generation interval than the symptomatic group [54]. Naturally, these differences are less pronounced under current conditions where the effective reproduction number is low and the entire population has been sheltering in place. It will be interesting to see if the effects of asymptomatic transmission become more visible as we gradually relax the current constraints and allow all individuals to move around and interact with others more freely. Seasonality, effects of different temperature and humidity, and other unknown factors may also influence the extent of transmission.

### Estimates of the infectious asymptomatic population may vary, but general trends are similar

Throughout the past months, an increasing number of researchers around the globe have started to characterize the size of the asymptomatic population to better understand the outbreak dynamics of COVID-19 [30]. The reported undercount varies largely with values on the order of one estimated by the World Health Organization [80] and values on the order of ten reported by the Center for Disease Control and Prevention [6]. Two major challenges drive the interest in these studies: estimating the severity of the outbreak, e.g., hospitalization and mortality rates [16], and predicting the success of surveillance and control efforts, e.g., contact tracing or vaccination [19]. This is especially challenging now-in almost complete lockdown-when the differences in transmission dynamics between the symptomatic and asymptomatic populations are small and difficult to quantify. However, as Figure 4 suggests, these transmission dynamics can have a significant effect on the size of the asymptomatic population: For infectious periods of *C*_a_ = 0.5, 1.0, and 2.0 *C*_s_, the maximum infectious population in Santa Clara County varies from 0.70% and 1.23% to 2.10%. Interestingly, not only the sum of the infectious and recovered populations, but also the uncertainty of their prediction, remain relatively insensitive to variations in the infectious period. To explore whether this is a universal trend, we perform the same analysis for nine different locations at which COVID-19 antibody prevalence was measured in a representative sample of the population, Heinsberg (NRW, Germany) [71], Ada County (ID, USA) [4], New York City (NY, USA) [58], Santa Clara County (CA, USA) [3], Denmark [14], Geneva Canton (Switzerland) [72], Netherlands [67], Rio Grande do Sul (Brasil) [66], and Belgium [64]. The fraction of the symptomatic population in these nine locations is *v*_s_ = 20.00%, 7.90%, 5.76%, 1.77%, 6.95%, 10.34%, 17.31%, 8.10%, and 10.21% respectively, broadly representing the range of reported symptomatic versus estimated total cases worldwide [3, 4, 9, 12, 14, 18, 24, 32, 34, 46, 58, 59, 64, 65, 68, 66, 67, 71, 72, 74, 76, 78, 81]. Of the nine locations we analyzed here, Heinsberg tested IgG and IgA, Ada County tested IgG, New York City tested IgG, Santa Clara County tested IgG and IgM, Denmark tested IgG and IgM, Geneva tested IgG, the Netherlands tested IgG, IgM and IgA, Rio Grande do Sul tested IgG and IgM, and Belgium tested IgG. While we did include reported uncertainty on the seroprevalence data, seroprevalence would likely have been higher if all locations had tested for all three antibodies. Despite these differences, the effective reproduction numbers *R*(*t*) and the infectious and recovered populations *I*_s_, *I*_a_, and *R* in Figure 6 display remarkably similar trends: In most locations, the effective reproduction number *R*(*t*) drops rapidly to values below one within a window of about three weeks after the lockdown date. However, the maximum infectious population, a value that is closely monitored by hospitals and health care systems, varies significantly ranging from 0.28% and 0.38% in Rio Grande do Sul and Denmark respectively to 3.54% and 6.11% in Heinsberg and New York City respectively. This is consistent with the reported ‘superspreader’ events in these last two locations. An effect that we do not explicitly address is that immune response not only results COVID-19 antibodies (humoral response), but also from innate and cellular immunity [23]. While it is difficult to measure the effects of the unreported asymptomatic group directly, and discriminate it precisely from innate and cellular immunity, mathematical models can provide valuable insight into how this population modulates the outbreak dynamics and the potential of successful outbreak control [40].

### Simulations provide a window into the outbreak date

Santa Clara County was home to the first individual who died with COVID-19 in the United States. Although this happened as early as February 6, the case remained unnoticed until April 22 [1]. The unexpected new finding suggests that the new coronavirus was circulating in the Bay Area as early as January. The estimated uncertainty on the exposed, symptomatic infectious, and asymptomatic infectious populations of our model allows us to estimate the initial outbreak date dates back to January 20 (Figure 7). This back-calculated early outbreak date is in line with our intuition that COVID-19 is often present in a population long before the first official case is reported. Interestingly, our analysis comes to this conclusion purely based on a local serology antibody study [3] and the number of reported cases after lockdown [63].

### Limitations

Our approach naturally builds in and learns several levels of uncertainties. By design, this allows us to estimate sensitivities and credible intervals for a number of important model parameters and discover important features and trends. Nevertheless, it has a few limitations, some of them by design, some simply limited by the current availability of data: First, our current SEIIR model assumes a similar contact rate *β*(*t*) for symptomatic and asymptomatic individuals. While we can easily adjust this in the model by defining individual symptomatic and asymptomatic rates *β_s_*(*t*) and >*β*_a_(*t*), we currently do not have data on the temporal evolution of the hidden asymptomatic infectious population *I*_a_(*t*) and longitudinal large population antibody follow-up studies would be needed to appropriately calibrate *β*_a_(*t*). Second, the ratio between the symptomatic and asymptomatic populations *v*_s_: *v*_a_ can vary over time, especially, as we have shown, if both groups display notably different dynamics, in our model represented through *C*_s_ and *C*_a_. Since this can have serious effects on the overall reproduction number *R*(*t*), and with it on required outbreak control strategies, it seems critical to perform more tests and learn the dynamics of the fractions *v*_s_(*t*) and *v*_a_(*t*) of both groups. Third, and this is not only true for our specific model, but for COVID-19 forecasts in general, all predictions can be sensitive to the amount of testing in time. As such, they crucially rely on testing policies and testing capacities. We expect to see a significant increase in the symptomatic-to-asymptomatic, or rather detected-to-undetected, ratio as we move towards systematically testing larger fractions of the population and more and more people who have no symptoms at all. The intensity of testing increases in most locations during our simulation period. For example, in Santa Clara County, testing was extremely limited until early April, increased substantially in the first three weeks of April, and even more after. Including limited testing and more undocumented cases during the early outbreak would shift the case distribution towards earlier days, and predict an even earlier outbreak date. When longitudinal antibody studies become available, additional methodologies can be developed to correct for this limitation which include a dynamic evolution for the symptomatic/asymptomatic fractions. Fourth, while we have included uncertainty in the seroprevalence data, the nine locations we analyzed here tested different types of antibodies and had different sampling procedures. Seroprevalence could have been higher if all locations had tested for the same three antibodies and data may differ depending on biases introduced by the sampling procedure. Finally, our current model does not explicitly account for innate and cellular immunity. If the fraction of the population with innate and cellular immunity is substantially high, we would anticipate a smaller susceptible population and a larger and earlier protective immunity overall. These, and other limitations related to the availability of information, can be easily addressed and embedded in our model and will naturally receive more clarification as studies and data become available in the coming months.

## 5. Conclusions

The rapid and devastating development of the COVID-19 pandemic has raised many open questions about its outbreak dynamics and unsuccessful outbreak control. From an outbreak management standpoint-in the absence of effective vaccination and treatment-the two most successful strategies in controlling an infectious disease are isolating infectious individuals and tracing and quarantining their contacts. Both critically rely on a rapid identification of infections, typically through clinical symptoms. Recent antibody prevalence studies could explain why these strategies have largely failed in containing the COVID-19 pandemic: Increasing evidence suggests that the number of unreported asymptomatic cases could outnumber the reported symptomatic cases by an order of magnitude or more. Mathematical modeling, in conjunction with reported symptomatic case data, antibody seroprevalence studies, and machine learning allows us to infer, in real time, the epidemiology characteristics of COVID-19. We can now visualize the invisible asymptomatic population, estimate its role in disease transmission, and quantify the confidence in these predictions. A better understanding of asymptomatic transmission will help us evaluate strategies to manage the impact of COVID-19 on both our economy and our health care system. A large asymptomatic population is associated with a high risk of community spread and could require a conscious shift from containment to mitigation induced by behavior changes. Our study suggests that, until vaccination and treatment become available, increasing population awareness, encouraging increased hygiene, mandating the use of face masks, restricting travel, and promoting physical distancing could be the most successful strategies to manage the impact of COVID-19 on both our economy and our health care system.

## Data Availability

All data are available on public servers and are cited in the references of the manuscript

## Appendix

To visualize the effect of asymptomatic transmission on the outbreak dynamics of COVID-19, we perform a sensitivity analysis with respect to the two most relevant parameters associated with asymptomatic transmission, the symptomatic fraction *v*_s_ and the asymptomatic infectious period *C*_a_. To highlight the effect of both parameters, we visualize the exposed, symptomatic and asymptomatic infectious, and recovered populations of our SEIIR model,

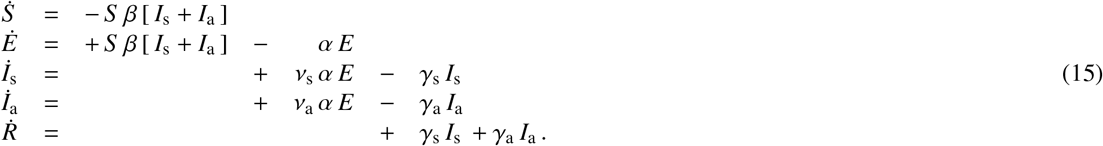

We assume that the symptomatic and asymptomatic contact rates are equal, *β = β*_s_ *= β*_a_, and that the differences between symptomatic and asymptomatic transmission manifest themselves exclusively in the infectious periods *C*_s_ = 1/*γ*_s_ and *C*_a_ = 1/*γ*_a_. For illustrative purposes, rather than using a dynamic contact rate of Gaussian random walk type as we have proposed in the main body of this manuscript, we adopt a smooth, monotonically decreasing dynamic contact rate of hyperbolic tangent type [43],

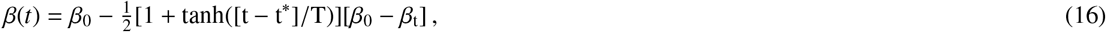

where *β*_0_ is the basic contact rate, *β*_t_ is the reduced contact rate, *t^*^* is the adaptation time, and *T* is the adaptation speed. To mimic realistic outbreak dynamics, we adopt the basic contact rate *β*_0_ *=* 0.65 /days, the reduced contact rate (*β*_t_ = 0.10 /days, the adaptation time *t*^*^ = 18.61 days, and the adaptation speed *T* = 10.82 /days from the mean values across the 27 countries of the European union [43]. Similar to the main body of this manuscript, we select a latent period of *A* = 1/*α* = 2.5 days and a symptomatic infectious period of *C*_a_ = 1/*γ*_a_ = 6.5 days. For the sensitivity analysis, we vary the symptomatic fraction *v*_s_ = [0.0,…, 1.0] at fixed *C*_a_ = 3.25 days and *C*_a_ = 13 days and the asymptomatic infectious period *C*_a_ = [3.25,…, 13.00] days at fixed *v*_s_ = 0.2 and *v*_s_ = 0.8. For a symptomatic fraction of *v*_s_ = 0.2, the undercount is four, i.e., for every reported case, there are four unreported cases. For a symptomatic fraction of *v*_s_ = 0.8, the undercount is one fourth, i.e., for every four reported cases, there is one unreported case.

Figure 8 highlights the effect of the symptomatic fraction *v*_s_. The dark red, orange, green, and blue curves correspond to the special case of the regular SEIR model (6) without asymptomatic transmission and thus *v*_s_ = 1.0 and *v*_a_ = 0.0. The lighter curves show the effect of decreasing the symptomatic fraction towards *v*_s_ = 0.0 and *v*_a_ = 1.0. Interestingly, the effect of asymptomatic transmission on the overall outbreak dynamics varies not only with the symptomatic fraction, but is also highly sensitive to the ratio of the symptomatic and asymptomatic infectious periods. For the smaller asymptomatic infectious period of *C*_a_ = 0.5 *C*_s_, the outbreak dynamics are governed by the orange symptomatic population *I*_s_ and the blue recovered population *R* decreases with increasing asymptomatic transmission *v*_a_. For the larger asymptomatic infectious period of *C*_a_ = 2 *C*_s_, the outbreak dynamics are governed by the green asymptomatic population *I*_a_ and the blue recovered population *R* increases with increasing asymptomatic transmission.

**Figure 8:**
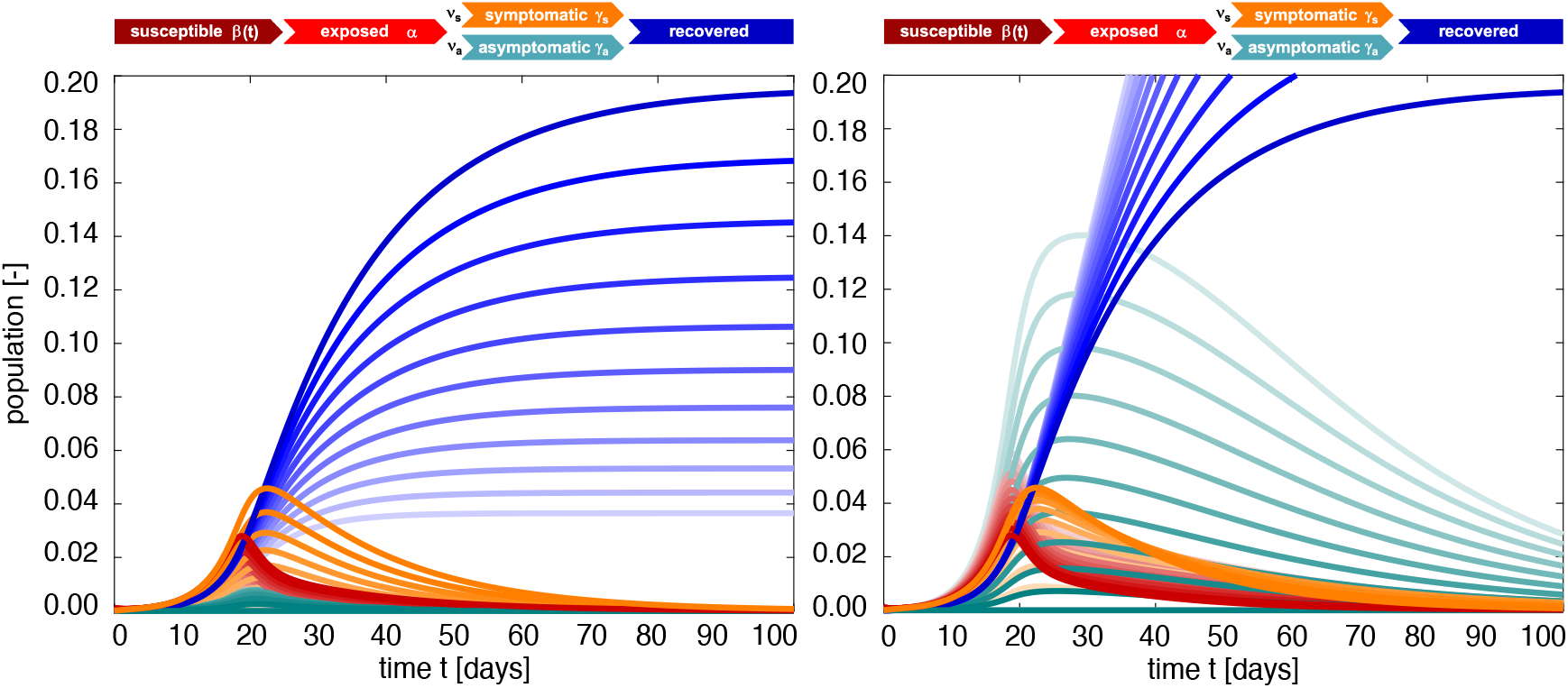
SEIIR epidemiology model. Sensitivity with respect to the symptomatic fraction *v*_s_. Darkest curves correspond to the special case of no asymptomatic transmission, *v*_s_ = 1.0, lightest curves to only asymptomatic transmission, *v*_s_ = 0.0. For a smaller asymptomatic infectious period of *C*_a_ = 0.5 *C*_s_, the outbreak dynamics are governed by the symptomatic population *I*_s_ and the recovered population *R* decreases with increasing asymptomatic transmission *v*_a_ (left). For a larger asymptomatic infectious period of *C*_a_ = 2 *C*_s_, the outbreak dynamics are governed by the asymptomatic population *I*_a_ and the recovered population increases with increasing asymptomatic transmission (right). Latent period *A =* 2.5 days, symptomatic infectious period *C*_s_ = 6.5 days, asymptomatic infectious period *C*_a_ = 3.25 days (left) and *C*_a_ = 13 days (right), and contact rate 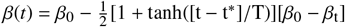 with *β*_0_ = 0.65 /days, *β*_t_ = 0.10 /days, *t*^*^ = 18.61 days, and *T* = 10.82 days.

Figure 9 shows the effect of the asymptomatic infectious period *C*_a_. The darkest red, orange, green, and blue curves correspond to a larger asymptomatic infectious period of *C*_a_ = 2 *C*_s_, the lightest curves to a smaller period *C*_a_ = 0.5 *C*_s_. The middle curves correspond to the special case for which the asymptomatic and symptomatic populations display the same dynamics, *C*_a_ = *C*_s_, such that the overall recovered population becomes independent of the symptomatic fraction. This implies that middle blue recovered populations of both graphs are identical and also identical to the dark blue curves of Figure 8. For the larger symptomatic fraction of *v*_s_ = 0.8, the outbreak dynamics are governed by the orange symptomatic population *I*_s_ and the blue recovered population *R* is relatively insensitive to the asymptomatic infectious period *C*_a_. For the smaller symptomatic fraction of *v*_s_ = 0.2, the outbreak dynamics are governed by the green asymptomatic population *I*_a_ and the blue recovered population *R* increases markedly with an increasing asymptomatic infectious period *C*_a_.

**Figure 9:**
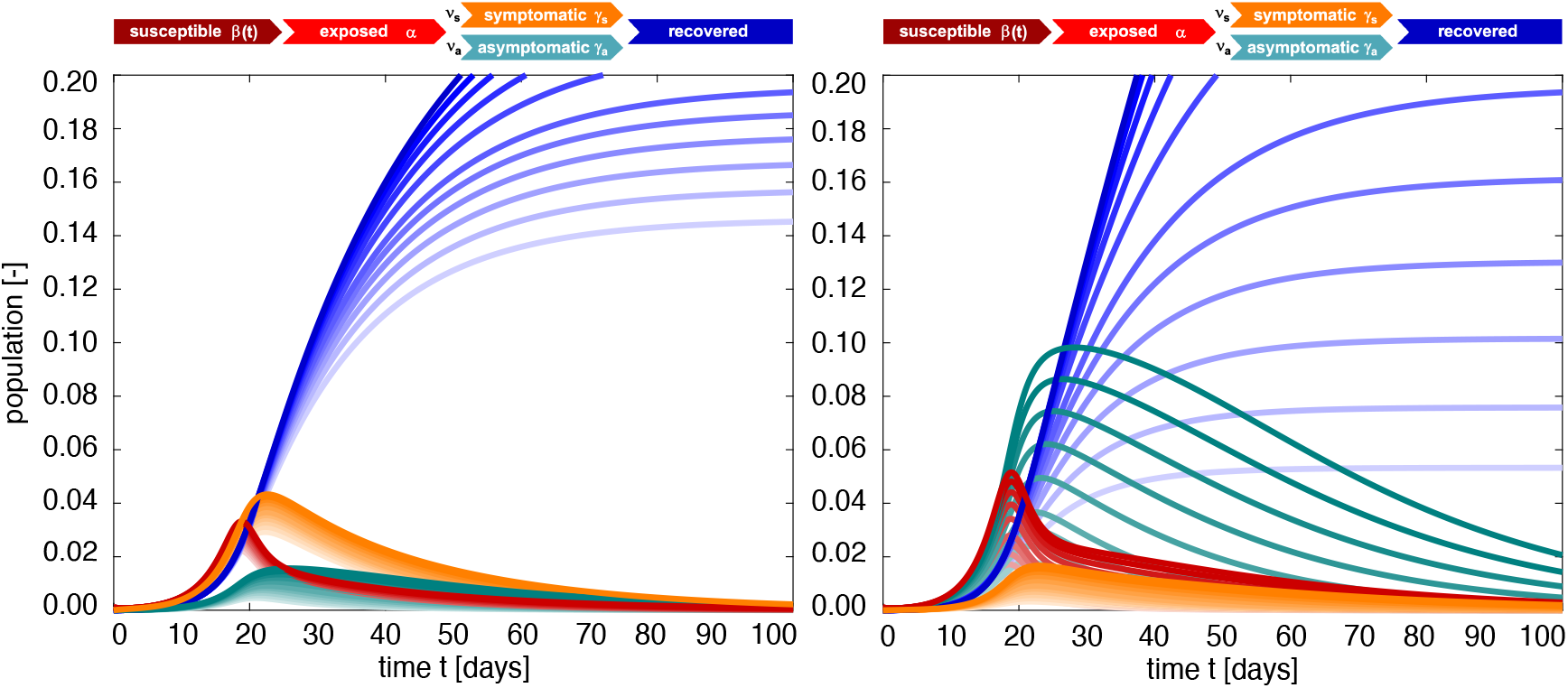
SEIIR epidemiology model. Sensitivity with respect to the asymptomatic infectious period *C*_a_. Darkest curves correspond to a larger asymptomatic infectious period of *C*_a_ = 2 *C*_s_, lightest curves to a smaller period *C*_a_ = 0.5 *C*_s_. For a larger symptomatic fraction of *v*_s_ = 0.8, the outbreak dynamics are governed by the symptomatic population *I*_s_ and the recovered population *R* is relatively insensitive to the asymptomatic infectious period *C*_a_ (left). For a smaller symptomatic fraction of *v*_s_ = 0.2, the outbreak dynamics are governed by the asymptomatic population *I*_a_ and the recovered population *R* increases markedly with an increasing asymptomatic infectious period *C*_a_ (right). Latent period *A* = 2.5 days, symptomatic infectious period *C*_s_ = 6.5 days, asymptomatic infectious period *C*_a_ = 3.25 days (left) and *C*_a_ = 13 days (right), and contact rate 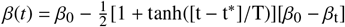 with *β*_0_ = 0.65 /days, *β_t_* = 0.10 /days, *t^*^* = 18.61 days, and *T* = 10.82 days.

Taken together, this sensitivity analysis shows that the outbreak dynamics of COVID-19 are highly sensitive to asymptomatic transmission. This sensitivity becomes more pronounced the more the asymptomatic infectious period differs from its symptomatic counterpart. This difference could be a natural result of asymptomatic individuals isolating themselves less since they are unaware that they can infect others. Knowing the precise dynamics of asymptomatic transmission is therefore critical for a successful management of COVID-19.

## Acknowledgements

This work was supported by a Stanford Bio-X IIP Seed Grant (M.P. and E.K.), by the NIH Grant U01 HL119578 (E.K.), by a DAAD Fellowship (K.L.), and by the Stanford COVID-19 Seroprevalence Study Fund (J.B. and E.B.).

## Notes

### Competing Interest Statement

The authors have declared no competing interest.

### Funding Statement

This work was supported by a DAAD Fellowship (K.L.), by a Stanford Bio-X IIP Seed Grant (M.P. and E.K.), and by the Stanford COVID-19 Seroprevalence Study Fund (J.B. and E.B.).

### Summary of Updates

Appendix added

